# Cost-Effectiveness of Screening Asymptomatic Carotid Stenosis by Atherosclerotic Cardiovascular Risk

**DOI:** 10.1101/2023.11.28.23299146

**Authors:** Jinyi Zhu, Janice Jhang, Hanxuan Yu, Alvin I Mushlin, Hooman Kamel, Nathaniel Alemayehu, John C Giardina, Ajay Gupta, Ankur Pandya

## Abstract

**Importance:** Extracranial internal carotid artery stenosis (50-99% arterial narrowing) is an important risk factor for ischemic stroke. Yet, the benefits and harms of targeted screening for asymptomatic carotid artery stenosis (ACAS) have not been assessed in population-based studies.

**Objective:** To estimate the cost-effectiveness of one-time, targeted ACAS screening stratified by atherosclerotic cardiovascular disease (ASCVD) risk using the American Heart Association’s Pooled Cohort Equations.

**Design, Setting, and Participants:** We developed a lifetime microsimulation model of ACAS and stroke for a hypothetical cohort representative of US adults aged 50–80 years without stroke history. We used the Cardiovascular Health Study to estimate the probability and severity of ACAS based on individual characteristics (e.g., age, sex, smoking status, blood pressure, and cholesterol). Stroke risks were functions of these characteristics and ACAS severity. In the model, individuals testing positive for >70% stenosis with Duplex ultrasound and a confirmatory diagnostic test undergo revascularization, which may reduce the risk of stroke but also introduces complication risks. Diagnostic performance parameters, revascularization benefits and risks, utility weights, and costs were estimated from published sources. Cost-effectiveness was assessed from the health care sector perspective using a $100,000/quality-adjusted life year (QALY) threshold.

**Main Outcomes and Measures:** Estimated stroke events prevented, lifetime costs, QALYs, and incremental cost-effectiveness ratios (ICERs) associated with ACAS screening. Costs (2023 USD) and QALYs were discounted at 3% annually.

**Results:** We found that screening individuals with a 10-year ASCVD risk >30% was the most cost-effective strategy, with an ICER of $89,000/QALY. This strategy would make approximately 11.9% of the population eligible for screening, averting an estimated 24,084 strokes. Results were sensitive to variations in the efficacy and complication risk of revascularization. In probabilistic sensitivity analysis, screening those in lower ASCVD risk groups (0–20%) only had a 0.6% chance of being cost-effective.

**Conclusion and Relevance:** A one-time screening may only be cost-effective for adults at a relatively high ASCVD risk. Our findings provide a framework that can be adapted as future clinical trial data continue to improve our understanding of the role of revascularization and intensive medical therapy in contemporary stroke prevention secondary to carotid disease.

## Introduction

Stroke is the 5th leading cause of mortality and a major cause of disability in the US.^1^ Of all strokes, 14% can be attributed to thromboembolism from previously asymptomatic stenosis (50-99%) of the extracranial internal carotid artery.^2^ Population-level screening with duplex ultrasonography (DUS) offers potential early intervention for those with asymptomatic carotid artery stenosis (ACAS), but this approach is controversial given evidence gaps regarding its effectiveness and risks.^2^

For patients with severe carotid artery stenosis, previous trials involving both symptomatic and asymptomatic patients have found benefits associated with revascularization procedures such as carotid endarterectomy (CEA) or carotid artery stenting (CAS).^3–5^ However, these procedures also carry a risk of complications, and the optimal approach for managing ACAS remains unclear. The Carotid Revascularization and Medical Management for Asymptomatic Carotid Stenosis Trial (CREST-2) is currently underway to evaluate the risks and benefits of these procedures compared to modern intensive medical management.^6^

Currently, the US Preventive Services Task Force (USPSTF) and most US professional societies recommend against universal ACAS screening.^7–10^ However, several societies recommend considering DUS screening and potential revascularization for asymptomatic patients with multiple stroke risk factors,^7,9,10^ and evidence suggests a substantial volume of screenings is still being performed in the US.^11–13^ Because several factors increase the risk for both carotid artery stenosis and ischemic stroke (e.g., older age, male sex, hypertension),^1,7^ targeted screening in high-risk subgroups could reduce the number needed to screen to prevent one stroke. The 2021 USPSTF Evidence Review highlighted the necessity for improved risk identification tools.^8^ In response to this need, we sought to estimate the cost-effectiveness of one-time, targeted ACAS screening of the US adult population, stratified by atherosclerotic cardiovascular disease (ASCVD) risk using the American Heart Association’s Pooled Cohort Equations (PCEs).^14^

## Methods

### Study Cohort

We developed a microsimulation model of ACAS and stroke for a hypothetical cohort representative of all US adults aged 50–80 years without prior transient ischemic attack (TIA) or stroke history (N=100,473,000).^1^ Model individuals were sampled by weights from the 2013-2014, 2015-2016, and 2017-2018 waves of the National Health and Nutrition Examination Survey (NHANES). Extracted characteristics included age, sex, race, history of diabetes, smoking status (current vs. any other), total and high-density lipoprotein cholesterol levels, systolic and diastolic blood pressure, history of ASCVD, and hypertension treatment. Since NHANES did not collect carotid artery stenosis status, we probabilistically assigned individuals to initial stenosis categories based on detailed methods described below.^15,16^

### Simulation Model

The model, outlined in Figure 1, tracks individuals’ annual progression or regression of carotid artery stenosis and the occurrences of TIA and stroke, while monitoring their health outcomes and associated costs over their lifetime. Pre-stroke natural history followed our previously published carotid stenosis model, which tracked stenosis progression, regression, and revascularization, and stroke incidence based on stenosis category; we added extensions incorporating ACAS screening and TIA incidence for this analysis.^17^ Acute and post-acute stroke events and outcomes were based on another published stroke model that we previously developed and validated.^18^ All individuals are subject to age- and sex-specific, non-stroke-related background mortality drawn from the US life tables.^19^

**Figure 1:**
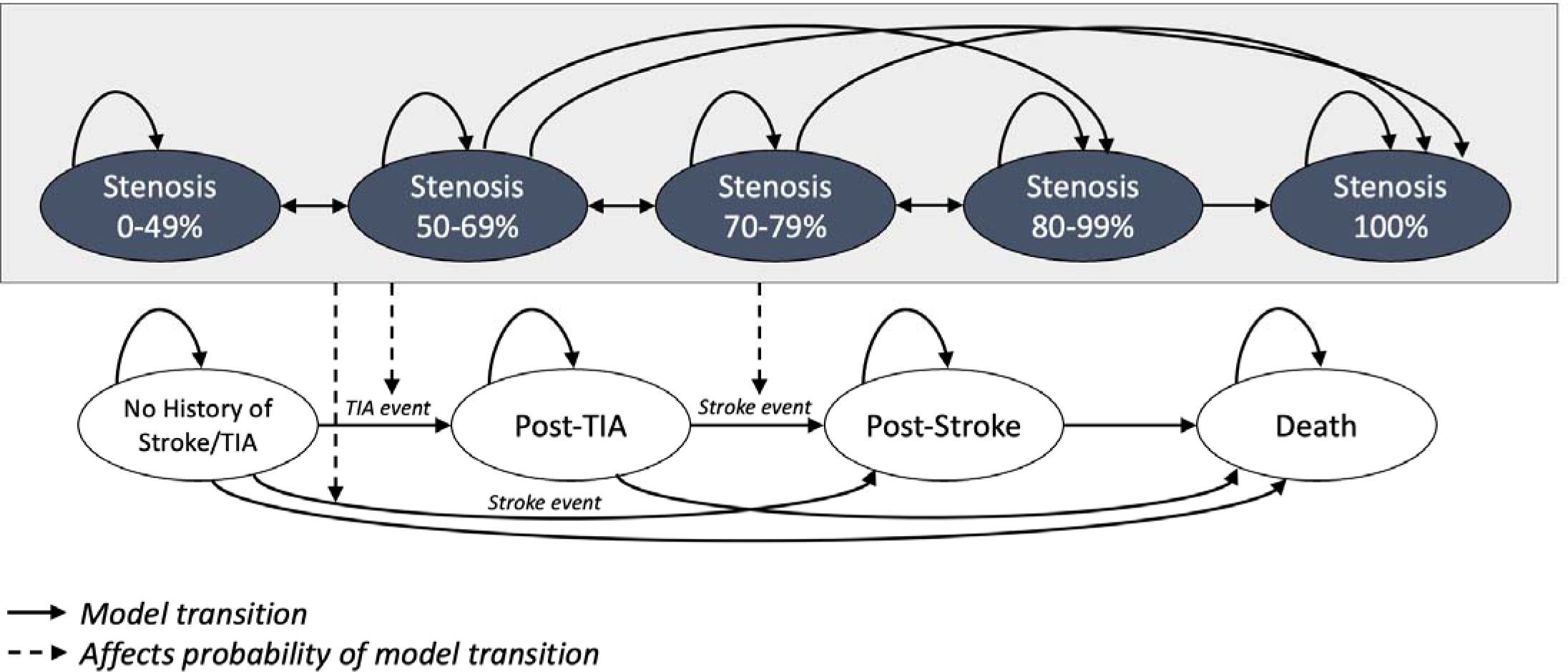
Model schematic. Individuals with no history of stroke can progress or regress in carotid artery stenosis severity. Severity of carotid artery stenosis affects individuals’ probability of experiencing a transient ischemic attack (TIA) or stroke. Following a revascularization procedure, individuals are categorized as having 0–49% carotid artery stenosis.

Each year, individuals in the 0–49% narrowing category may progress to a 50–69% stenosis state, while those with 50–99% stenosis may advance up to three categories or regress by one annually. We assumed that patients with 100% stenosis cannot regress and are ineligible for revascularization. A higher stenosis severity is associated with an increased annual risk of TIA and stroke. Individuals’ risk of TIA is based on their age, sex, and stenosis severity.^20^ Any individual who experiences a TIA receives a confirmatory diagnostic test of computed tomography angiography or magnetic resonance angiography in the same year and undergoes revascularization if the confirmatory test reveals carotid stenosis.

Once an individual develops a stroke event, they transition into our previously published acute and post-acute stroke model.^18^ In brief, patients’ acute stroke outcomes are governed by their resulting modified Rankin Scale score (mRS, a discrete score between 0 and 6, where mRS=0 indicates no symptoms and mRS=6 indicates death). A more severe mRS is assigned a higher recurrent stroke risk, higher mortality, lower utility weight, and higher annual post-stroke management costs for stroke survivors. See Zhu et al.^18^ for further details on the acute and post-acute stroke model structure and inputs.

### Model Parameterization

We derived parameters, including event rates, probabilities, test characteristics, intervention effects, utility weights, and costs, from several prospective cohort studies and existing literature (detailed in Table 1). Specifically, to determine the baseline probability of moderate-to-severe ACAS (≥50% stenosis) for each individual, we first created a multivariate logistic regression model from the Cardiovascular Health Study (CHS) and validated this prediction model using a split-sample approach in a test set of 30% of the data. This model was then calibrated to the sex- and age-specific prevalence of ACAS from published sources.^15^ Individuals predicted to have stenosis ≥50% were then randomly assigned to 1 of the 4 stenosis blockage categories (50–69%, 70–79%, 80–99%, or 100%) with weights obtained through calibration to data reported in Hirt 2014.^16^

**Table 1:**
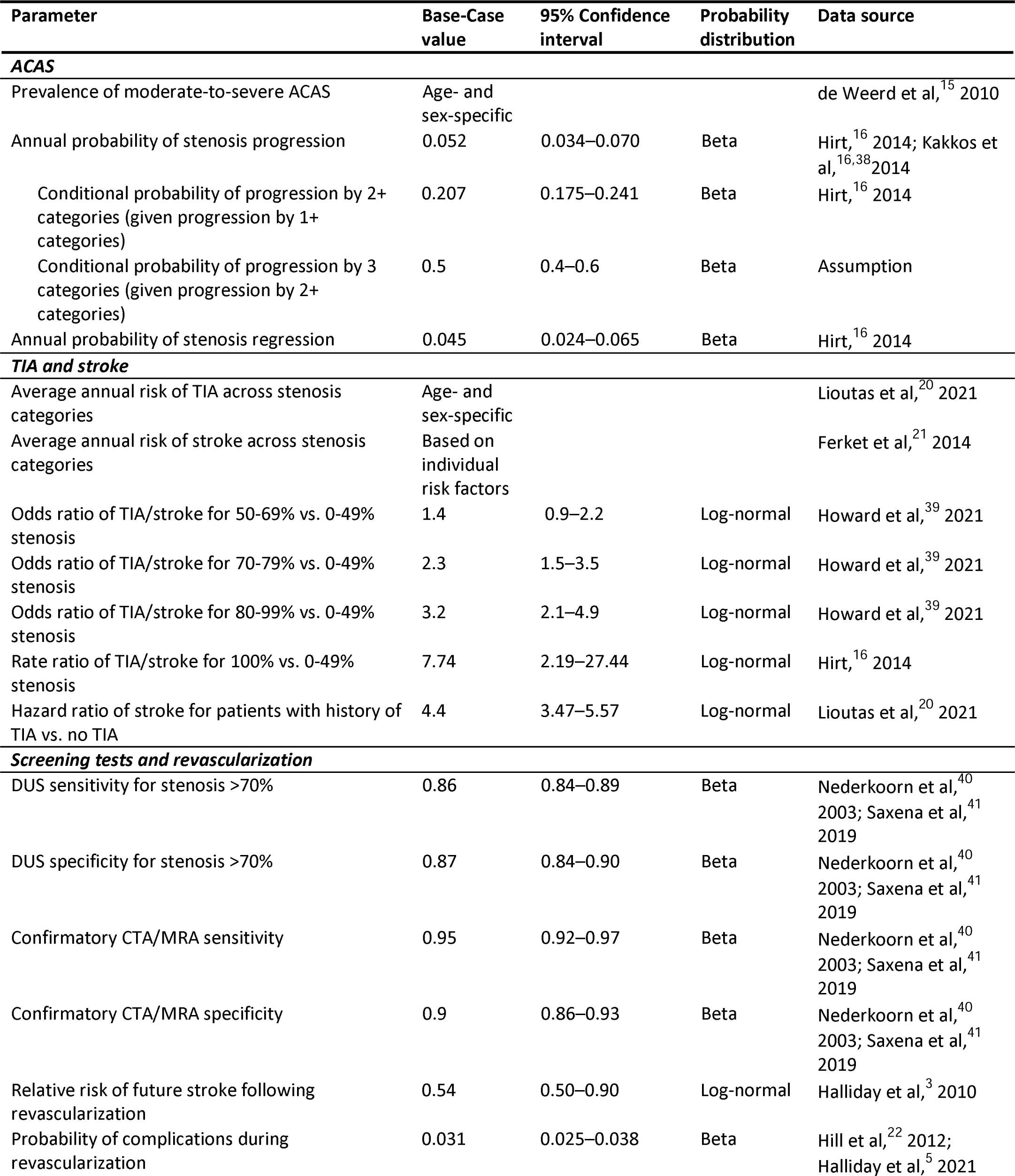
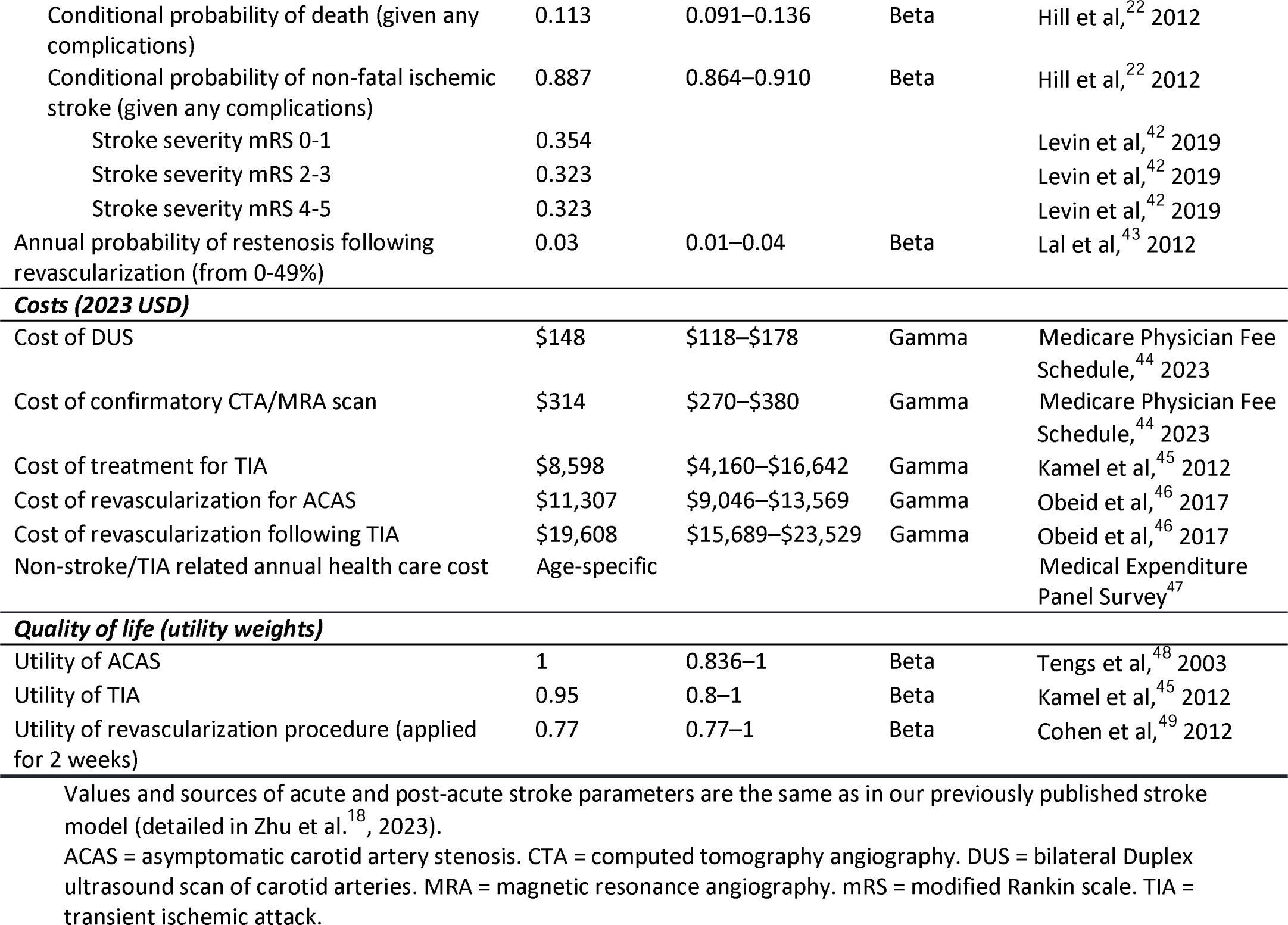
Parameter inputs.

| Parameter | Base-Case value | 95% Confidence interval | Probability distribution | Data source |
| --- | --- | --- | --- | --- |
| **ACAS** |  |  |  |  |
| Prevalence of moderate-to-severe ACAS | Age- and sex-specific |  |  | de Weerd et al,<sup>15</sup> 2010 |
| Annual probability of stenosis progression | 0.052 | 0.034–0.070 | Beta | Hirt,<sup>16</sup> 2014; Kakkos et al,<sup>16,38</sup> 2014 |
| Conditional probability of progression by 2+ categories (given progression by 1+ categories) | 0.207 | 0.175–0.241 | Beta | Hirt,<sup>16</sup> 2014 |
| Conditional probability of progression by 3 categories (given progression by 2+ categories) | 0.5 | 0.4–0.6 | Beta | Assumption |
| Annual probability of stenosis regression | 0.045 | 0.024–0.065 | Beta | Hirt,<sup>16</sup> 2014 |
| **TIA and stroke** |  |  |  |  |
| Average annual risk of TIA across stenosis categories | Age- and sex-specific |  |  | Lioutas et al,<sup>20</sup> 2021 |
| Average annual risk of stroke across stenosis categories | Based on individual risk factors |  |  | Ferket et al,<sup>21</sup> 2014 |
| Odds ratio of TIA/stroke for 50-69% vs. 0-49% stenosis | 1.4 | 0.9–2.2 | Log-normal | Howard et al,<sup>39</sup> 2021 |
| Odds ratio of TIA/stroke for 70-79% vs. 0-49% stenosis | 2.3 | 1.5–3.5 | Log-normal | Howard et al,<sup>39</sup> 2021 |
| Odds ratio of TIA/stroke for 80-99% vs. 0-49% stenosis | 3.2 | 2.1–4.9 | Log-normal | Howard et al,<sup>39</sup> 2021 |
| Rate ratio of TIA/stroke for 100% vs. 0-49% stenosis | 7.74 | 2.19–27.44 | Log-normal | Hirt,<sup>16</sup> 2014 |
| Hazard ratio of stroke for patients with history of TIA vs. no TIA | 4.4 | 3.47–5.57 | Log-normal | Lioutas et al,<sup>20</sup> 2021 |
| **Screening tests and revascularization** |  |  |  |  |
| DUS sensitivity for stenosis >70% | 0.86 | 0.84–0.89 | Beta | Nederkoorn et al,<sup>40</sup> 2003; Saxena et al,<sup>41</sup> 2019 |
| DUS specificity for stenosis >70% | 0.87 | 0.84–0.90 | Beta | Nederkoorn et al,<sup>40</sup> 2003; Saxena et al,<sup>41</sup> 2019 |
| Confirmatory CTA/MRA sensitivity | 0.95 | 0.92–0.97 | Beta | Nederkoorn et al,<sup>40</sup> 2003; Saxena et al,<sup>41</sup> 2019 |
| Confirmatory CTA/MRA specificity | 0.9 | 0.86–0.93 | Beta | Nederkoorn et al,<sup>40</sup> 2003; Saxena et al,<sup>41</sup> 2019 |
| Relative risk of future stroke following revascularization | 0.54 | 0.50–0.90 | Log-normal | Halliday et al,<sup>3</sup> 2010 |
| Probability of complications during revascularization | 0.031 | 0.025–0.038 | Beta | Hill et al,<sup>22</sup> 2012; Halliday et al,<sup>5</sup> 2021 |
| Conditional probability of death (given any complications) | 0.113 | 0.091–0.136 | Beta | Hill et al,<sup>22</sup> 2012 |
| Conditional probability of non-fatal ischemic stroke (given any complications) | 0.887 | 0.864–0.910 | Beta | Hill et al,<sup>22</sup> 2012 |
| Stroke severity mRS 0-1 | 0.354 |  |  | Levin et al,<sup>42</sup> 2019 |
| Stroke severity mRS 2-3 | 0.323 |  |  | Levin et al,<sup>42</sup> 2019 |
| Stroke severity mRS 4-5 | 0.323 |  |  | Levin et al,<sup>42</sup> 2019 |
| Annual probability of restenosis following revascularization (from 0-49%) | 0.03 | 0.01–0.04 | Beta | Lal et al,<sup>43</sup> 2012 |
| **Costs (2023 USD)** |  |  |  |  |
| Cost of DUS | \$148 | \$118–\$178 | Gamma | Medicare Physician Fee Schedule,<sup>44</sup> 2023 |
| Cost of confirmatory CTA/MRA scan | \$314 | \$270–\$380 | Gamma | Medicare Physician Fee Schedule,<sup>44</sup> 2023 |
| Cost of treatment for TIA | \$8,598 | \$4,160–\$16,642 | Gamma | Kamel et al,<sup>45</sup> 2012 |
| Cost of revascularization for ACAS | \$11,307 | \$9,046–\$13,569 | Gamma | Obeid et al,<sup>46</sup> 2017 |
| Cost of revascularization following TIA | \$19,608 | \$15,689–\$23,529 | Gamma | Obeid et al,<sup>46</sup> 2017 |
| Non-stroke/TIA related annual health care cost | Age-specific |  |  | Medical Expenditure Panel Survey<sup>47</sup> |
| **Quality of life (utility weights)** |  |  |  |  |
| Utility of ACAS | 1 | 0.836–1 | Beta | Tengs et al,<sup>48</sup> 2003 |
| Utility of TIA | 0.95 | 0.8–1 | Beta | Kamel et al,<sup>45</sup> 2012 |
| Utility of revascularization procedure (applied for 2 weeks) | 0.77 | 0.77–1 | Beta | Cohen et al,<sup>49</sup> 2012 |

Values and sources of acute and post-acute stroke parameters are the same as in our previously published stroke model (detailed in Zhu et al.<sup>18</sup>, 2023). ACAS = asymptomatic carotid artery stenosis. CTA = computed tomography angiography. DUS = bilateral Duplex ultrasound scan of carotid arteries. MRA = magnetic resonance angiography. mRS = modified Rankin scale. TIA = transient ischemic attack.

Ischemic stroke risk was calculated using 10-year cumulative incidence functions developed by Ferket et al.^21^ Factors used to predict individual risk of ischemic stroke included age, sex, race, current smoking status, diabetes, antihypertensive medication use, systolic blood pressure, and history of coronary heart disease.^21^ To capture the longitudinal development of stroke risk, we utilized 3 datasets (the CHS, the Atherosclerosis Risk in Communities Study, and the Multi-Ethnic Study of Atherosclerosis) in a two-step approach: First, we calculated each individual’s 10-year predicted ischemic stroke risk at each follow-up measurement, employing the equations by Ferket et al.^21^ Subsequently, we applied a linear mixed effects model, relating this 10-year risk to follow-up time and the interaction between time and baseline stroke risk factors. Estimates from this longitudinal model were then incorporated into the simulation model to update stroke risk every 10 years. History of TIA, stenosis severity, and history of revascularization affected individuals’ stroke risk using separate relative risk estimates that were applied multiplicatively (see Table 1).

### Screening Strategies

In our model, a specific subset of individuals receives a one-time ACAS screening, the selection of which depends on the strategy being evaluated and each individual’s risk of ASCVD. In addition to screening none and screening all, we evaluated 7 ACAS screening strategies that were specified based on 10-year ASCVD risk thresholds: >35%, >30%, >25%, >20%, >15%, >10%, and >5%. Individuals’ 10-year ASCVD risks were predicted from the PCEs, using their age, sex, race, diabetes, smoking, total and high-density lipoprotein cholesterol, systolic blood pressure, and blood pressure treatment.^14^

ACAS screening is defined as a two-stage sequence of tests, starting with a DUS of carotid arteries, followed by a confirmatory test of computed tomography angiography or magnetic resonance angiography if the DUS indicates 70–99% stenosis in either carotid artery. If both tests are positive, individuals then undergo a revascularization procedure (CEA or CAS), in a similar fashion to the recommended guidelines for symptomatic patients.^7,9^ CEA and CAS are associated with complication risks (stroke or death).^5,22^ After successful revascularization, individuals’ level of stenosis is categorized as 0–49% (which reduces their risk). However, given their prior history of stenosis, they experience an elevated risk of stroke compared to those without a revascularization history in the same stenosis category. On average, revascularization reduces the incidence of non-perioperative stroke by 46% in our base case based on results from the Asymptomatic Carotid Surgery Trial (ACST-1),^3^ and we varied this assumption in sensitivity analyses.

### Model Outcomes

Our primary health outcomes were the number of strokes averted and quality-adjusted life-years (QALYs) gained relative to no ACAS screening over the lifetimes of the model cohort. Quality of life was quantified using utility weights for post-TIA and post-stroke health states and a disutility weight for revascularization procedures. Total lifetime costs include the costs of TIA and acute and post-acute stroke care, costs of screening tests and revascularization, and usual health care costs for individuals without a history of stroke. Costs were assessed from a health care sector perspective and inflation-adjusted to 2023 US dollars.^23^ We calculated the incremental cost-effectiveness ratio (ICER) as the additional cost per QALY gained for screening strategies that were not dominated (i.e., more expensive and less effective relative to another strategy). We used a base-case cost-effectiveness threshold of $100,000/QALY.^24^ Future health and cost outcomes were discounted annually at 3%.^25^

### Sensitivity Analyses

We performed a set of univariate sensitivity analyses, varying each model parameter between its 95% confidence intervals or plausible ranges while holding the other parameters constant. We also conducted a two-way sensitivity analysis on two key parameters with large uncertainty and evidence gaps: the efficacy (i.e., relative risk of future non-perioperative stroke) and complication risk of revascularization for individuals with ACAS.

In probabilistic sensitivity analyses, we varied model input values based on pre-specified probability distributions and estimated the probability of each strategy being cost-effective at various cost-effectiveness thresholds. The parameter ranges and distributions used in our sensitivity analyses are summarized in Table 1. We report the mean estimates for each model outcome from 1,000 samples of parameter sets from the probabilistic sensitivity analysis as our base-case results, alongside the 95% uncertainty intervals (UIs).

We performed all statistical analyses using R (v4.2.2) and programmed the simulation model using the Rcpp package (v1.0.10). We followed the Consolidated Health Economic Evaluation Reporting Standards guideline in reporting our study (Appendix Table 1).^26^

## Results

### Model Validation

Summary statistics of our simulated model cohort, stratified by 10-year ASCVD risk, are presented in Appendix Table 2. An increased ASCVD risk was correlated with a higher proportion of men, older age, and greater prevalence of ACAS. Our ACAS prediction model derived from CHS data showed acceptable discrimination (c-statistic=0.72 in the test set), similar to the performance reported by Poorthuis et al. (c-statistic=0.75).^27^ Our calibrated age- and sex-stratified ACAS prevalence estimates were comparable to data reported in a meta-analysis of population-based studies (Appendix Table 3).^15^

**Table 2:** Base-case cost-effectiveness results.

|  | Proportion of US population screened | Relative to screen none |  |  | Relative to the next less intensive screening strategy (i.e., compared to the strategy immediately above) |  |  |
| --- | --- | --- | --- | --- | --- | --- | --- |
| | | Number of strokes averted | Cost (\$, in millions) | QALYs (in thousands) | Incremental Cost, (\$, in millions) | Incremental QALYs (in thousands) | ICER (\$/QALY) |
| Screen none | 0% | Reference |  |  |  |  |  |
| Screen 10yr ASCVD Risk >35% | 7.5% | 18,243<br>(17,112 to 19,493) | 2,000<br>(1,604 to 2,337) | 55.7<br>(50.5 to 61.9) | 2,000<br>(1,604 to 2,337) | 55.7<br>(50.5 to 61.9) | 36,000<br>(26,000 to 45,000) |
| Screen 10yr ASCVD Risk >30% | 11.9% | 24,084<br>(22,420 to 25,895) | 3,473<br>(2,851 to 3,994) | 72.2<br>(64 to 81.3) | 1,473<br>(1,256 to 1,655) | 16.5<br>(13.7 to 19.7) | 89,000<br>(65,000 to 119,000) |
| Screen 10yr ASCVD Risk >25% | 16.9% | 29,610<br>(27,448 to 32,008) | 5,171<br>(4,297 to 5,906) | 85.4<br>(74 to 98.3) | 1,699<br>(1,442 to 1,918) | 13.2<br>(10.1 to 17) | 129,000<br>(84,000 to 181,000) |
| Screen 10yr ASCVD Risk >20% | 24.8% | 37,312<br>(34,283 to 40,798) | 7,994<br>(6,706 to 9,068) | 103.2<br>(86.5 to 122.6) | 2,823<br>(2,408 to 3,149) | 17.7<br>(12.2 to 24.1) | 159,000<br>(96,000 to 225,000) |
| Screen 10yr ASCVD Risk >15% | 36.3% | 44,241<br>(39,871 to 49,180) | 12,252<br>(10,379 to 13,800) | 107.3<br>(82.7 to 136.2) | 4,258<br>(3,674 to 4,758) | 4.2<br>(-3.2 to 14.1) | 1,022,000<br>(461,400 to dominated) |
| Screen 10yr ASCVD Risk >10% | 52.3% | 48,820<br>(43,099 to 55,726) | 18,423<br>(15,672 to 20,618) | 88.6<br>(54.7 to 131.5) | Dominated |  |  |
| Screen 10yr ASCVD Risk >5% | 77.8% | 47,405<br>(39,614 to 57,147) | 29,096<br>(25,086 to 32,361) | 6.8<br>(-43.6 to 72.3) | Dominated |  |  |
| Screen All | 100% | 40,053<br>(30,634 to 52,243) | 38,748<br>(33,509 to 43,061) | -87.1<br>(-154 to 1.8) | Dominated |  |  |

Intervals in parentheses denote 95% uncertainty intervals. All results are reported for the US adult population aged 50–80 years (total N = 100,473,000). Both costs and QALYs are discounted at 3% per year. Costs are reported in 2023 USD. ASCVD = atherosclerotic cardiovascular disease. ICER = incremental cost-effectiveness ratio. QALY = quality-adjusted life-years

### Base-Case Results

The mean values and 95% UIs of all population-level outcomes of each screening strategy are reported in Table 3, while individual-level (i.e., per-person) results are shown in Appendix Table 4. Implementing a more lenient screening threshold (i.e., lower ASCVD risk threshold) would consistently result in higher total discounted lifetime costs. Meanwhile, the number of strokes averted would be maximized at a 10-year ASCVD risk threshold of >10%, and total discounted lifetime QALYs were maximized at a threshold of >15%. This non-monotonicity in health outcomes across screening thresholds is explained by the dual effect of ACAS screening: it diagnoses more cases of moderate-to-severe ACAS for early stroke prevention, but simultaneously increases mortality and morbidity due to a larger number of complications from revascularization procedures.

Under our base-case $100,000/QALY cost-effectiveness threshold, the optimal strategy would be screening adults aged 50–80 years who have a 10-year ASCVD risk greater than 30%. This strategy yielded an ICER of $89,000/QALY (95% UI, $65,000–119,000/QALY), compared to screening those with an ASCVD risk greater than 35%. Under this screening strategy, 11.9% of the population was estimated to be eligible for ACAS screening, which could avert 24,084 (22,420–25,895) strokes over the cohort’s lifetime relative to no screening.

### Sensitivity Analyses

When each parameter of interest was individually adjusted within its confidence interval or plausible range, the optimal screening strategy was sensitive to variation in 14 parameters (Figure 2), including stenosis progression and regression rates, performance and cost of screening tests, revascularization efficacy and complication rate, probability of post-revascularization restenosis, and recurrent stroke risk. Varying any other model parameter alone did not affect the optimal screening strategy.

**Figure 2:**
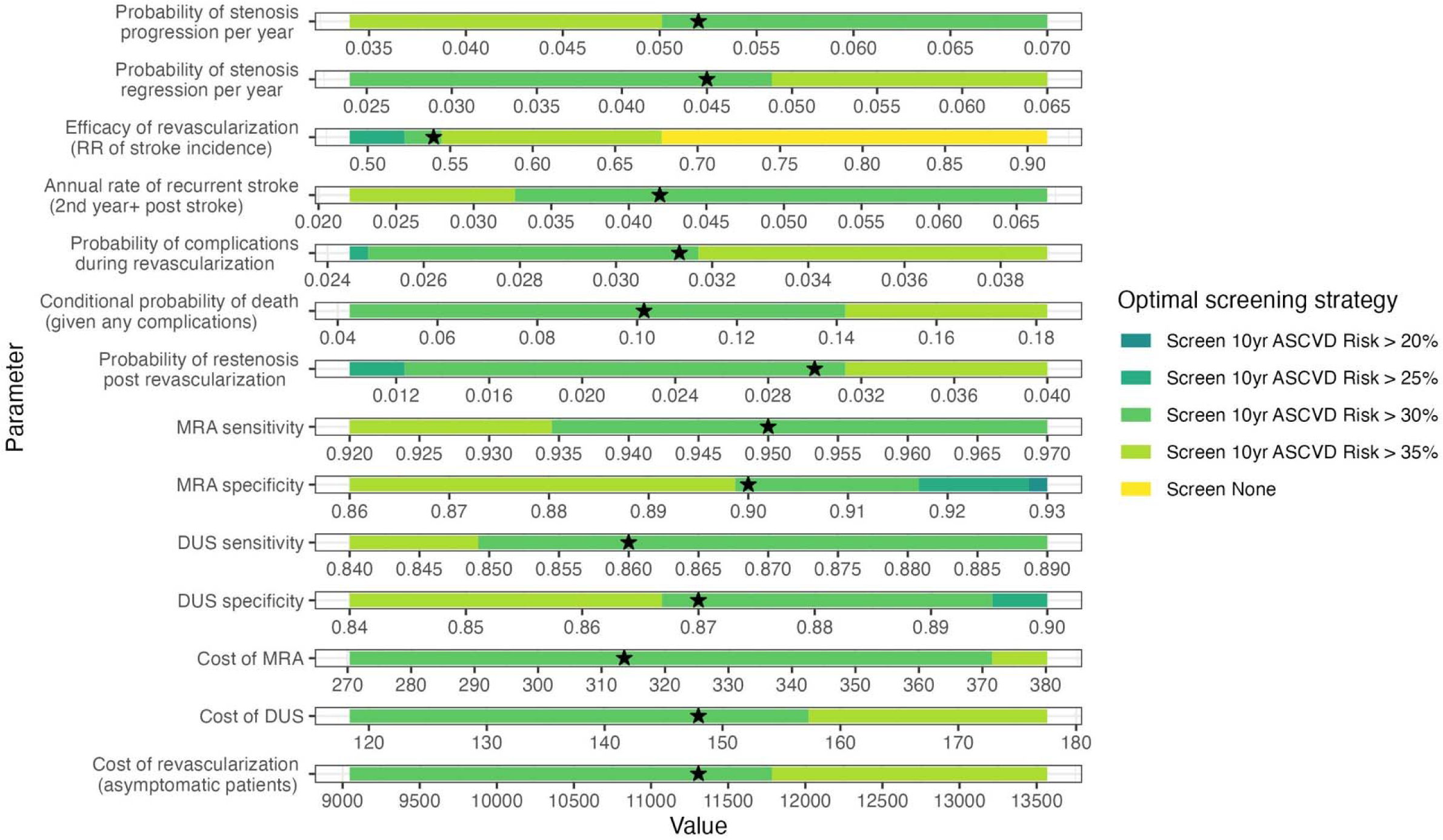
One-way sensitivity analysis results. Stars denote the base-case parameter values. The graph displays the 14 parameters that significantly influenced the optimal screening strategy at the $100,000/quality-adjusted life year threshold, as indicated by sensitivity to variations within these parameters. The optimal screening strategy did not change while varying any of the other model parameters. ASCVD = atherosclerotic cardiovascular disease. DUS = bilateral Duplex ultrasound scan of carotid arteries. MRA = magnetic resonance angiography. RR = relative risk.

Figure 3 shows the two-way sensitivity analysis results, focusing on the efficacy and complication risk of revascularization procedures. We found that ACAS screening would be less favorable with a higher probability of complications or worse efficacy of revascularization than our base-case assumptions. Nonetheless, adopting a higher ASCVD risk threshold for screening (i.e., >35%) could remain cost-effective in some of those scenarios.

**Figure 3:**
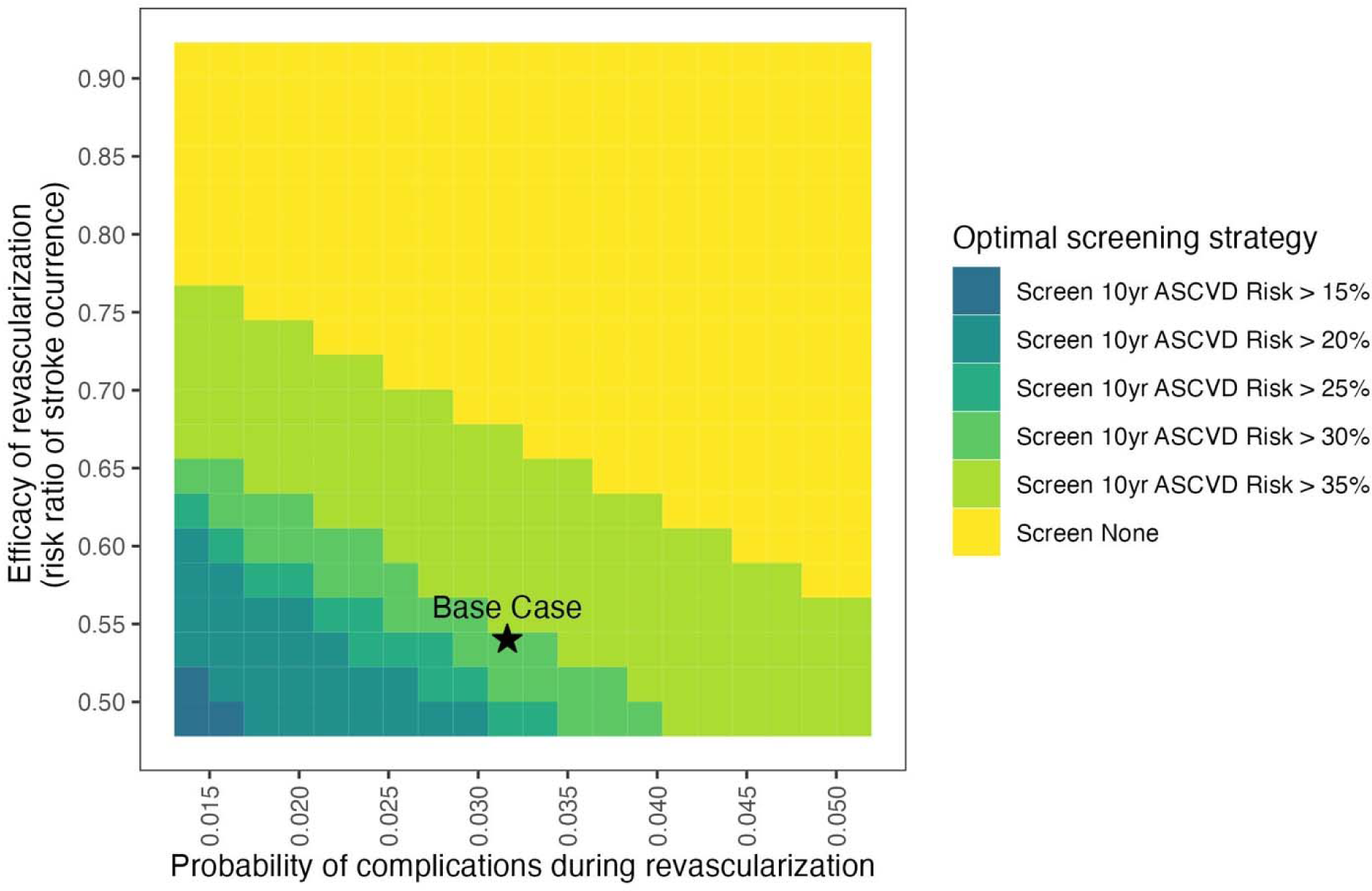
Impact of revascularization complications and efficacy on the optimal screening strategy. This heat map represents the results of a two-way sensitivity analysis evaluating how variations in the probability of revascularization complications and the efficacy of revascularization affect the choice of optimal screening strategy at the $100,000/quality-adjusted life-year threshold. ASCVD = atherosclerotic cardiovascular disease.

Probabilistic sensitivity analysis results are shown in cost-effectiveness acceptability curves and frontier (Appendix Figure 1A), where we varied all model parameters at the same time. For a range of cost-effectiveness thresholds from $0–200,000 per QALY, the acceptability curves show the probabilities of each ASCVD risk-based strategy being cost-effective, while the frontier displays the cost-effective strategy on average. We found large uncertainty in the optimal screening threshold at the base-case $100,000/QALY threshold: Using >35%, >30%, 25%, and >20% as screening thresholds had probabilities of 36.0%, 27.8%, 12.0%, and 22.9%, respectively, of being optimal across 1,000 parameter sets. This uncertainty was largely driven by the uncertainty in the cost-effectiveness of screening for the 20–35% 10-year ASCVD risk groups (Appendix Figure 1B). Our analysis robustly showed that screening individuals in the >35% risk group would be cost-effective in 99.3% of the 1,000 iterations. Conversely, screening lower ASCVD risk groups (0–20%) was almost never cost-effective, as evidenced in only 0.6% of the iterations.

## Discussion

Our model-based cost-effectiveness analysis of targeted screening strategies for ACAS showed that screening individuals with 10-year ASCVD risk greater than 30% would meet conventional US standards for cost-effectiveness (<$100,000/QALY). This group comprises 11.9% of the US population aged 50–80 years. Most of the gains in stroke prevention would come from screening those at the highest ASCVD risk (>35%).

These results were sensitive to key assumptions regarding the effectiveness and safety of revascularization procedures. Model parameters were based on results from the Carotid Revascularization Endarterectomy Versus Stent Trial (CREST) and the first and second Asymptomatic Carotid Surgery Trials (ACST-1 and ACST-2).^3–5^ These trials did not compare the efficacy of revascularization plus modern intensive medical management to medical management alone. With modern medical advancements, particularly increasing utilization of anti-hypertensive and lipid-lowering medications, the benefits of revascularization may be diminishing.^28^ CREST-2 is underway to re-assess the relative benefit of revascularization compared to intensive medical management alone for ACAS. Once CREST-2 concludes, the results from our analysis can help translate the trial results into cost-effective screening recommendations. For example, our sensitivity analyses suggest that a relative risk of future stroke incidence following revascularization above 0.7 would result in no screening becoming the optimal strategy (Figure 2).

Our study directly responds to a critical need highlighted by the 2021 USPSTF Recommendation Statement.^8^ The USPSTF currently recommends against universal screening for ACAS, supported by most specialty societies.^7–10^ However, guidelines from the Society for Vascular Surgery^9^ and joint guidelines from multiple societies^10^ include exceptions for individuals with risk factors for stroke, which are present in nearly 1 in 3 US adults.^1^ Previous US-based cost-effectiveness analyses of ACAS screening, conducted before 2000, found that a one-time screening could be cost-effective if implemented in populations with a high prevalence of carotid stenosis or if revascularization procedures were performed by surgeons with low perioperative stroke and death rates.^29–31^ More recent studies have assessed more sophisticated risk stratification tools, e.g., the Carotid Mortality Index^32^ or ultrasound imaging to assess cerebrovascular reserve^17^ and plaque echolucency.^33^ However, no modern studies have assessed the value of targeted screening for ACAS. Our study innovates by using individual-level data to predict individual-specific risks of ACAS and stroke, incorporating results from more modern trials, and stratifying screening strategies based on individual ASCVD risk levels.

For risk stratification, we applied 10-year ASCVD risk estimates from the PCEs. The PCEs are commonly used in clinical practice and would be efficient to implement for potential ACAS screening decisions. Poorthuis et al. investigated the detection rate of ACAS through selective screening by 10-year ASCVD risk thresholds and demonstrated that selective screening could reduce the number needed to screen compared to population-wide screening.^34^ Our analysis complements these findings by explicitly identifying the optimal ASCVD risk threshold for selective screening. In November 2023, the American Heart Association introduced the PREVENT (Predicting Risk of Cardiovascular Disease EVENTs) risk prediction equations, which estimate both short- and long-term ASCVD risk while integrating broader health determinants and excluding race as a factor.^35^ Should the PREVENT equations start to be adopted in clinical practice, risk stratification for ACAS screening using the PREVENT equations rather than the PCEs could be evaluated in future cost-effectiveness studies.

Our study has several limitations. First, the NHANES data used to construct the model population did not report individuals’ carotid stenosis status, so we developed a multivariate logistic regression model to probabilistically determine the presence of ACAS. We validated our ACAS prediction model using a split sample approach and found acceptable but imperfect discrimination (c-statistic = 0.72). Our ACAS prediction model was based on the best model^36^ determined by Poorthuis et al.^27^ in an external validation study of proposed models to predict ACAS and included all overlapping covariates available in NHANES. That study found that most models proposed for ACAS prediction have modest discrimination; however, they can reliably identify subgroups at high risk of carotid stenosis, which could substantially reduce the number needed to screen to detect ACAS.^27^ Second, our study did not account for secular trends in population stroke risk over time, potentially underestimating the ongoing improvements in medical management of stroke risks.^37^ Nonetheless, if CREST-2 finds that the benefit of intensive medical management is comparable to revascularization, a targeted screening strategy may still be valuable to identify individuals for whom the potential benefits of an aggressive medical therapeutic regimen outweigh side effects. Finally, while our full simulation model has not been externally validated, key parts have been validated, including the previously published acute and post-acute stroke models.^18^ We also performed split-sample validation of individual-level prediction of moderate-to-severe carotid stenosis, and the predicted ACAS prevalence was well-calibrated to population-based studies.^15^

Based on current evidence, we estimate that ACAS screening at a relatively high 10-year predicted ASCVD risk (>30%) could be cost-effective. If CREST-2 confirms a relative stroke risk post-revascularization above 0.7, it might shift the balance against any screening. While our results are sensitive to key parameters that will be clarified in the CREST-2 trial, our sensitivity analysis results are structured to allow adaptation to future trial findings. In the interim, our findings can provide a basis for current clinical practice and policy decisions.

## Data Availability

All data produced in the present study are available upon reasonable request to the authors

## Author Contributions

Study concept and design: Zhu, Jhang, Yu, Mushlin, Kamel, Gupta, Pandya.

Acquisition, analysis, or interpretation of data: All authors.

Drafting of the manuscript: Zhu, Jhang, Yu, Pandya.

Critical revision of the manuscript for important intellectual content: All authors.

Statistical analysis: Zhu, Jhang, Yu, Alemayehu.

Obtained funding: Pandya.

Administrative, technical, or material support: Zhu, Pandya.

Study supervision: Zhu, Pandya.

## References

1. Tsao CW, Aday AW, Almarzooq ZI, et al. Heart Disease and Stroke Statistics-2023 Update: A Report From the American Heart Association. Circulation. 2023;147(8):e93-e621. doi:10.1161/CIR.0000000000001123

2. Naylor AR. Why is the management of asymptomatic carotid disease so controversial? The Surgeon. 2015;13(1):34–43. doi:10.1016/j.surge.2014.08.004

3. Halliday A, Harrison M, Hayter E, et al. 10-year stroke prevention after successful carotid endarterectomy for asymptomatic stenosis (ACST-1): a multicentre randomised trial. The Lancet. 2010;376(9746):1074–1084. doi:10.1016/S0140-6736(10)61197-X

4. Brott TG, Hobson RW, Howard G, et al. Stenting versus Endarterectomy for Treatment of Carotid-Artery Stenosis. N Engl J Med. 2010;363(1):11–23. doi:10.1056/NEJMoa0912321

5. Halliday A, Bulbulia R, Bonati LH, et al. Second asymptomatic carotid surgery trial (ACST-2): a randomised comparison of carotid artery stenting versus carotid endarterectomy. The Lancet. 2021;398(10305):1065–1073. doi:10.1016/S0140-6736(21)01910-3

6. Mott M, Koroshetz W, Wright CB. CREST-2: Identifying the Best Method of Stroke Prevention for Carotid Artery Stenosis. Stroke. 2017;48(5):e130–e131. doi:10.1161/STROKEAHA.117.016051

7. Kleindorfer DO, Towfighi A, Chaturvedi S, et al. 2021 Guideline for the Prevention of Stroke in Patients With Stroke and Transient Ischemic Attack: A Guideline From the American Heart Association/American Stroke Association. Stroke. 2021;52(7):e364-e467. doi:10.1161/STR.0000000000000375

8. US Preventive Services Task Force. Screening for Asymptomatic Carotid Artery Stenosis: US Preventive Services Task Force Recommendation Statement. JAMA. 2021;325(5):476–481. doi:10.1001/jama.2020.26988

9. AbuRahma AF, Avgerinos ED, Chang RW, et al. Society for Vascular Surgery clinical practice guidelines for management of extracranial cerebrovascular disease. J Vasc Surg. 2022;75(1, Supplement):4S–22S. doi:10.1016/j.jvs.2021.04.073

10. Brott TG, Halperin JL, Abbara S, et al. 2011 ASA/ACCF/AHA/AANN/AANS/ACR/ASNR/CNS/SAIP/SCAI/SIR/SNIS/SVM/SVS guideline on the management of patients with extracranial carotid and vertebral artery disease. Stroke. 2011;42(8):e464–540. doi:10.1161/STR.0b013e3182112cc2

11. Radomski TR, Zhao X, Lovelace EZ, et al. Use and Cost of Low-Value Health Services Delivered or Paid for by the Veterans Health Administration. JAMA Intern Med. 2022;182(8):832–839. doi:10.1001/jamainternmed.2022.2482

12. Anderson TS, Leonard S, Zhang AJ, et al. Trends in Low-Value Carotid Imaging in the Veterans Health Administration From 2007 to 2016. JAMA Netw Open. 2020;3(9):e2015250. doi:10.1001/jamanetworkopen.2020.15250

13. Boudreau E, Schwartz R, Schwartz AL, et al. Comparison of Low-Value Services Among Medicare Advantage and Traditional Medicare Beneficiaries. JAMA Health Forum. 2022;3(9):e222935. doi:10.1001/jamahealthforum.2022.2935

14. Goff DC, Lloyd-Jones DM, Bennett G, et al. 2013 ACC/AHA Guideline on the Assessment of Cardiovascular Risk. Circulation. 2014;129(25_suppl_2):S49–S73. doi:10.1161/01.cir.0000437741.48606.98

15. de Weerd M, Greving JP, Hedblad B, et al. Prevalence of asymptomatic carotid artery stenosis in the general population: an individual participant data meta-analysis. Stroke. 2010;41(6):1294–1297. doi:10.1161/STROKEAHA.110.581058

16. Hirt LS. Progression rate and ipsilateral neurological events in asymptomatic carotid stenosis. Stroke. 2014;45(3):702–706. doi:10.1161/STROKEAHA.111.613711

17. Pandya A, Gupta A, Kamel H, Navi BB, Sanelli PC, Schackman BR. Carotid Artery Stenosis: Cost-effectiveness of Assessment of Cerebrovascular Reserve to Guide Treatment of Asymptomatic Patients. Radiology. 2015;274(2):455–463. doi:10.1148/radiol.14140501

18. Zhu J, Kamel H, Gupta A, et al. Prioritizing Quality Measures in Acute Stroke Carel]: A Cost-Effectiveness Analysis. Ann Intern Med. 2023;176(5):649–657. doi:10.7326/M22-3186

19. Arias E, Xu J. United States Life Tables, 2018. Natl Vital Stat Rep Cent Dis Control Prev Natl Cent Health Stat Natl Vital Stat Syst. 2020;69(12):1–45.

20. Lioutas VA, Ivan CS, Himali JJ, et al. Incidence of Transient Ischemic Attack and Association With Long-term Risk of Stroke. JAMA. 2021;325(4):373–381. doi:10.1001/jama.2020.25071

21. Ferket BS, Kempen BJH van, Wieberdink RG, et al. Separate prediction of intracerebral hemorrhage and ischemic stroke. Neurology. 2014;82(20):1804–1812. doi:10.1212/WNL.0000000000000427

22. Hill MD, Brooks W, Mackey A, et al. Stroke after carotid stenting and endarterectomy in the Carotid Revascularization Endarterectomy versus Stenting Trial (CREST). Circulation. 2012;126(25):3054–3061. doi:10.1161/CIRCULATIONAHA.112.120030

23. U.S. Bureau of Economic Analysis. BEA Interactive Data Application, “Table 1.1.3. Real Gross Domestic Product, Quantity Indexes.” Published 2023. Accessed January 21, 2024. https://apps.bea.gov/iTable/?reqid=19&step=2%23reqid%3D19&step=2&isuri=1&1921=survey#eyJhcHBpZCI6MTksInN0ZXBzIjpbMSwyLDNdLCJkYXRhIjpbWyJDYXRlZ29yaWVzIiwiU3VydmV5Il0sWyJOSVBBX1RhYmxlX0xpc3QiLCIzIl1dfQ==

24. Neumann PJ, Cohen JT, Weinstein MC. Updating Cost-Effectiveness — The Curious Resilience of the $50,000-per-QALY Threshold. N Engl J Med. 2014;371(9):796–797. doi:10.1056/NEJMp1405158

25. Sanders GD, Neumann PJ, Basu A, et al. Recommendations for Conduct, Methodological Practices, and Reporting of Cost-effectiveness Analyses: Second Panel on Cost-Effectiveness in Health and Medicine. JAMA. 2016;316(10):1093–1103. doi:10.1001/jama.2016.12195

26. Husereau D, Drummond M, Augustovski F, et al. Consolidated Health Economic Evaluation Reporting Standards 2022 (CHEERS 2022) Statement: Updated Reporting Guidance for Health Economic Evaluations. Value Health. 2022;25(1):3–9. doi:10.1016/j.jval.2021.11.1351

27. Poorthuis MHF, Halliday A, Massa MS, et al. Validation of Risk Prediction Models to Detect Asymptomatic Carotid Stenosis. J Am Heart Assoc. 2020;9(8):e014766. doi:10.1161/JAHA.119.014766

28. Matyori A, Brown CP, Ali A, Sherbeny F. Statins utilization trends and expenditures in the U.S. before and after the implementation of the 2013 ACC/AHA guidelines. Saudi Pharm J SPJ. 2023;31(6):795–800. doi:10.1016/j.jsps.2023.04.002

29. Derdeyn CP, Powers WJ. Cost-effectiveness of screening for asymptomatic carotid atherosclerotic disease. Stroke. 1996;27(11):1944–1950. doi:10.1161/01.str.27.11.1944

30. Lee TT, Solomon NA, Heidenreich PA, Oehlert J, Garber AM. Cost-Effectiveness of Screening for Carotid Stenosis in Asymptomatic Persons. Ann Intern Med. 1997;126(5):337–346. doi:10.7326/0003-4819-126-5-199703010-00001

31. Yin D, Carpenter JP. Cost-effectiveness of screening for asymptomatic carotid stenosis. J Vasc Surg. 1998;27(2):245–255. doi:10.1016/s0741-5214(98)70355-6

32. Keyhani S, Madden E, Cheng EM, et al. Risk Prediction Tools to Improve Patient Selection for Carotid Endarterectomy Among Patients With Asymptomatic Carotid Stenosis. JAMA Surg. 2019;154(4):336–344. doi:10.1001/jamasurg.2018.5119

33. Baradaran H, Gupta A, Anzai Y, Mushlin AI, Kamel H, Pandya A. Cost Effectiveness of Assessing Ultrasound Plaque Characteristics to Risk Stratify Asymptomatic Patients With Carotid Stenosis. J Am Heart Assoc. 2019;8(21):e012739. doi:10.1161/JAHA.119.012739

34. Poorthuis MHF, Sherliker P, de Borst GJ, et al. Detection rates of asymptomatic carotid artery stenosis and atrial fibrillation by selective screening of patients without cardiovascular disease. Int J Cardiol. 2023;391:131262. doi:10.1016/j.ijcard.2023.131262

35. Khan SS, Coresh J, Pencina MJ, et al. Novel Prediction Equations for Absolute Risk Assessment of Total Cardiovascular Disease Incorporating Cardiovascular-Kidney-Metabolic Health: A Scientific Statement From the American Heart Association. Circulation. 0(0). doi:10.1161/CIR.0000000000001191

36. de Weerd M, Greving JP, Hedblad B, et al. Prediction of asymptomatic carotid artery stenosis in the general population: identification of high-risk groups. Stroke. 2014;45(8):2366–2371. doi:10.1161/STROKEAHA.114.005145

37. Hackam DG. Optimal Medical Management of Asymptomatic Carotid Stenosis. Stroke. 2021;52(6):2191–2198. doi:10.1161/STROKEAHA.120.033994

38. Kakkos SK, Nicolaides AN, Charalambous I, et al. Predictors and clinical significance of progression or regression of asymptomatic carotid stenosis. J Vasc Surg. 2014;59(4):956–967.e1. doi:10.1016/j.jvs.2013.10.073

39. Howard DPJ, Gaziano L, Rothwell PM, Oxford Vascular Study. Risk of stroke in relation to degree of asymptomatic carotid stenosis: a population-based cohort study, systematic review, and meta-analysis. Lancet Neurol. 2021;20(3):193–202. doi:10.1016/S1474-4422(20)30484-1

40. Nederkoorn PJ, van der Graaf Y, Hunink MGM. Duplex ultrasound and magnetic resonance angiography compared with digital subtraction angiography in carotid artery stenosis: a systematic review. Stroke. 2003;34(5):1324–1332. doi:10.1161/01.STR.0000068367.08991.A2

41. Saxena A, Ng EYK, Lim ST. Imaging modalities to diagnose carotid artery stenosis: progress and prospect. Biomed Eng OnLine. 2019;18:66. doi:10.1186/s12938-019-0685-7

42. Levin SR, Farber A, Cheng TW, et al. Most patients experiencing 30-day postoperative stroke after carotid endarterectomy will initially experience disability. J Vasc Surg. 2019;70(5):1499–1505.e1. doi:10.1016/j.jvs.2019.02.035

43. Lal BK, Beach KW, Roubin GS, et al. Restenosis after carotid artery stenting and endarterectomy: a secondary analysis of CREST, a randomised controlled trial. Lancet Neurol. 2012;11(9):755–763. doi:10.1016/S1474-4422(12)70159-X

44. Search the Physician Fee Schedule | CMS. Published 2023. Accessed December 7, 2023. https://www.cms.gov/medicare/physician-fee-schedule/search

45. Kamel H, Johnston SC, Easton JD, Kim AS. Cost-effectiveness of dabigatran compared with warfarin for stroke prevention in patients with atrial fibrillation and prior stroke or transient ischemic attack. Stroke. 2012;43(3):881–883. doi:10.1161/STROKEAHA.111.641027

46. Obeid T, Alshaikh H, Nejim B, Arhuidese I, Locham S, Malas M. Fixed and variable cost of carotid endarterectomy and stenting in the United States: A comparative study. J Vasc Surg. 2017;65(5):1398–1406.e1. doi:10.1016/j.jvs.2016.11.062

47. MEPS-HC Data Tools – Medical Expenditure Panel Survey (MEPS) Household Component (HC). Published 2021. Accessed January 21, 2024. https://datatools.ahrq.gov/meps-hc/

48. Tengs TO, Lin TH. A meta-analysis of quality-of-life estimates for stroke. PharmacoEconomics. 2003;21(3):191–200. doi:10.2165/00019053-200321030-00004

49. Cohen DJ, Lavelle TA, Van Hout B, et al. Economic outcomes of percutaneous coronary intervention with drug-eluting stents versus bypass surgery for patients with left main or three-vessel coronary artery disease: one-year results from the SYNTAX trial. Catheter Cardiovasc Interv Off J Soc Card Angiogr Interv. 2012;79(2):198–209. doi:10.1002/ccd.23147

